# Programmed death ligand-1 and Programmed cell death protein-1 expression across the anal disease continuum and association with improved survival in anal cancer

**DOI:** 10.1101/2024.12.10.24318219

**Authors:** S. Chowdhury, C. Gasper, A. A. Lazar, K. Allaire, T. M. Darragh, L. Fong, J. M. Palefsky

**Affiliations:** Division of Infectious Diseases, Department of Medicine, University of California, San Francisco, CA, USA; Department of Pathology, University of California, San Francisco, CA, USA; Department of Epidemiology and Biostatistics, University of California, San Francisco, CA, USA; Division of Hematology/Oncology, Department of Medicine, University of California, San Francisco, CA, USA; Immunotherapy Integrated Research Center, Fred Hutchinson Cancer Center, Seattle, WA, USA

## Abstract

High-risk human papillomavirus is associated with anal high-grade intraepithelial lesion (aHSIL) and anal squamous cell carcinoma (aSCC). The prognostic significance of PD-L1 expression in aSCC and its impact on overall survival (OS) is controversial. ASCC can evade immune surveillance by co-opting the PD-L1/PD-1 immune checkpoint pathway, enhancing tumorigenesis. To assess the potential role of the PD-L1/PD-1 axis on tumor progression, we assessed PD-L1 and PD-1 expression on epithelial cells (ECs) and immune cells (ICs) by immunohistochemistry in benign anal tissue (n=22), aHSIL (n=22), and aSCC (n=52) from HIV-negative participants and people living with HIV. PD-L1 expression on EC was restricted to tumor cells with no expression in benign and HSIL tissues. PD-1 expression on ICs increased along the disease continuum from benign to SCC. The combined PD-L1 expression score on ECs and ICs showed a substantial increase from benign to aHSIL to aSCC. The combined positive score (CPS) for aSCC was 8.2. PD-L1 expression on IC in aSCC was more prominent than in tumor cells which correlated with increased IC infiltration and interferon-gamma secretion. 92% of aSCC demonstrated an adaptive PD-L1 expression pattern. HIV status did not affect PD-L1/PD-1 expression in benign, aHSIL or aSCC. PD-L1 expression in treatment naïve aSCC was associated with improved OS. Those with CPS of 0 had a higher risk of death [Hazard ratio 15.2 (95% CI: 3.3-69, p=0.0004; log-rank p<0.0001)] compared to those with CPS > 0. CPS may indicate the presence of immune activation and serve as a potential prognostic marker.

**Significance:** PD-L1 expression becomes more prominent as HPV-infected anal epithelial tissues progress from pre-cancer to cancer. ASCCs with high PD-L1/PD-1 expression indicates a reactive tumor microenvironment, making them promising candidates for immunotherapy.

## Introduction

Although a rare malignancy in the general population, the incidence of anal cancer is increasing both in the USA and globally. It is especially high in people living with human immunodeficiency virus (HIV) (PLWH). Anal cancer is now one of the leading non-AIDs-defining cancers in PLWH in the combined antiretroviral therapy (cART) era (1). Infection with one or more of fifteen high-risk (HR) HPVs is a major risk factor for the development of anal squamous cell carcinoma (aSCC) and its precursor lesion, anal high-grade intraepithelial lesion (aHSIL). HPV 16 is the most common oncogenic type found in aSCC (2). In addition to HPV and HIV, other risk factors include engaging in anal intercourse, history of HPV-associated vulvar or cervical cancer, immunosuppression due to organ transplantation and cigarette smoking (3).

To control HPV infection, the host mounts an innate immune response, consisting of innate immune cells (macrophages, natural killer (NK) cells, dendritic cells and neutrophils) and proinflammatory cytokines (IL-1, IL-6 and TNF-α ) (4). Interferons are another key antiviral defense mechanism that possess antiviral, immunoregulatory and antitumor effects, inhibiting HPV replication (5). The virus is often able to evade innate immunity by downregulating interferon secretion and response, thus delaying the activation of adaptive immunity. Even when adaptive immunity is mounted, orchestrated by T-helper cells (Th cells, CD4+) and cytotoxic T-cells (CTL, CD8+) the virus possesses mechanisms that render the cell-mediated response ineffective. This results in persistent viral infection (6).

HPV oncoproteins E6 and E7 play a major role in driving HPV-associated cancers and bind with high affinity to host cellular proteins p53 and pRb, respectively, in infected keratinocytes. Disruption of cell-cycle checkpoints leads to continuous cell proliferation and interference with various cellular processes including DNA repair, resulting in the accumulation of mutations in the host genome. Collectively, viral persistence, accumulation of cellular genetic alterations and viral immune evasion strategies allow for the development of HSIL and the progression of HSIL to cancer. Tumor-infiltrating lymphocytes (TILS) consisting of T-cells, B-cells, neutrophils, macrophages, eosinophils, basophils, and NK cells are recruited into the tumor microenvironment (TME) due to the expression of oncoproteins E6 and E7 (7). This recruitment promotes a chronic proinflammatory response that facilitates tumor progression (8). The rate of malignant transformation is notably higher among immunocompromised individuals (9). In PLWH, the likelihood of aHSIL progressing to cancer may be increased due to HIV-associated inflammation and chronic immune exhaustion (10).

Cancers often exploit various cellular pathways to evade immune recognition, including the exploitation of inhibitory checkpoints that terminate immune responses post-antigen activation. One such pathway involves upregulation of the checkpoint inhibitor molecule programmed death-ligand 1 (PD-L1). This strategy is employed in many HPV-associated cancers (cervical, anal, head and neck, vulvar, penile) dampening the host’s antigenic T cell response (11). PD-L1, found on tumor cells, parenchymal cells, endothelial cells, and macrophages, binds to programmed cell death protein 1 (PD-1) expressed on T cells, thus preventing T-cell-mediated immunosurveillance [11], facilitating an immunosuppressive microenvironment and tumor progression. The expression of PD-L1 is regulated by several biological mechanisms, its presence or absence therefore indicates different functional and clinical significance. PD-L1 positivity may be a result of genetic events leading to constitutive PD-L1 expression on cancer cells. Alternatively, inducible or adaptive PD-L1 expression on cancer and non-cancer cells occurs in response to interferon-gamma (IFN-γ) secretion by TILS (12).

Based on the presence or absence of PD-L1 expression and TILs the TME is classified into four types: adaptive immune resistance or inducible (PD-L1 positive/TILs present), immune ignorance (PD-L1 negative/TILs absent), intrinsic induction or constitutive (PD-L1 positive/TILs absent) and immune tolerance (PD-L1 negative/TILs present). This classification of TME may be useful in interpreting PD-L1 assay results and in predicting the efficacy of immune checkpoint inhibitor (ICI) treatment (12, 13).

Pharmacological interventions, including ICIs such as pembrolizumab, nivolumab and atezolizumab, block the PD-1 cell surface receptor or its ligand PD-L1, thereby restoring tumor antigen–specific T cell activity that was previously inhibited by the PD-1–PD-L1 interaction. In anal cancer, the use of PD-L1/ PD-1 monoclonal antibodies either alone or in combination with chemoradiotherapy (CRT) has shown promise in the treatment of metastatic and recurrent aSCC (14). While PD-L1/PD-1 therapy has demonstrated success in a subset of metastatic aSCC patients, its efficacy remains limited (15).

To address the limitations of ICIs, a comprehensive understanding of the functional role of PD-L1/PD-1 in aSCC pathogenesis is imperative. This necessitates characterization of PD-L1 and PD-1 expressing cells, along with elucidating the intricate crosstalk between these cells and other cellular components within the anal TME. Understanding the functionality of PD-L1/PD-1 in aHSIL is also essential to provide insight into the role of these proteins in progression to cancer. Several studies have tried to answer these questions in the context of aSCC but have been hindered by small sample size and divergent findings regarding PD-L1 expression patterns. These conflicting results have led to PD-L1 expression being viewed as either a positive, negative or inconsequential prognostic biomarker for aSCC. Furthermore, only one study has investigated the expression of PD-L1 and PD-1 in aHSIL.

We sought to bridge this gap by investigating the expression patterns of PD-L1 and PD-1 in epithelial and ICs in benign anal epithelium, aHSIL, and aSCC, in both PLWH and HIV-negative participants. Furthermore, we investigated the correlation between PD-L1 expression with clinical and prognostic factors to elucidate the clinical significance of the PD-L1/PD-1 axis in anal cancer.

## Materials and Methods

### Tissue collection

Archived formalin-fixed paraffin-embedded (FFPE) anal tissues (HSIL and SCC) were retrieved from the UCSF Department of Pathology. All blocks were de-identified of protected health information and were analyzed with institutional review board (IRB) permission. The FFPE samples consisted of biopsies collected in outpatient clinics and surgical specimens. Benign anal tissue specimens were obtained from patients undergoing anal surgery for treatment of aHSIL after obtaining their consent. Prior to electrocautery ablation of aHSIL, the surgeon biopsied peri-anal or intra-anal areas that clinically appeared to be normal. Additional samples of clinically normal anal tissue were obtained from patients undergoing surgery for anal fistulas and hemorrhoids. These were confirmed on histopathological examination to be benign. The tumor samples included biopsies, excisions and resections. All tumors used for the study were primary tumors. One sample per patient was collected. If a sample exhibited multiple histopathological grades, the more severe diagnostic category was considered for analysis.

### Patient characteristics

Anal tissues were collected between 2001–2021. All cases were reviewed in the electronic medical record until May 2023 and clinicopathological characteristics, including age, sex, race/ethnicity, HIV status, HIV viral load, and CD4+ count was collected. For aSCC specimens, location of biopsy or resection, tumor stage, tumor grade and whether patients underwent prior treatment was also collected. Prior treatment was defined as previous chemotherapy, radiation therapy or resection for any malignancy. The cohort was divided into three age groups 25-49, 50-69 and above 70 years of age. HIV viral load and CD4 values were obtained within 6 months of the date of diagnosis. HIV viral load reportable range was 20 to 10,000,000 copies/mL and it was categorized as undetectable (ND), >20 copies/mL and <1000 copies/mL, >1000 copies/mL. CD4 counts were divided into three groups: below 200 cells/mm^3^, 200-500 cells/mm^3^ and >500 cells/mm^3^. The patients were categorized based on their immune status into three groups: PLWH, HIV-negative immunosuppressed individuals (e.g., those who have undergone organ transplantation or have an autoimmune disease), and non-immunosuppressed individuals.

The analysis of outcomes included overall survival (OS), cancer-specific survival, disease progression and progression-free survival (PFS). Overall survival was defined as the duration in time from the date of diagnosis to death or last follow-up, with no restriction on the cause of death. Deaths attributed to cancer were those resulting from the spread of the disease or complications from treatments like chemotherapy or radiation, such as sepsis. Fatalities linked to HIV status but not directly caused by aSCC were classified as overall deaths but not included in the count of cancer-related deaths. Disease progression was defined as the time from date of diagnosis to persistent illness, local recurrence, the emergence of positive lymph nodes indicating the spread of the disease, and distant metastasis, whichever occurred first. We used data from radiological reports (CT scans, PET scans, and/or MRIs) and clinical follow-up notes to track disease progression. PFS was defined as the duration from diagnosis to death or progression, whichever occurred first. Survivors were censored at the time of the last documented follow-up.

### Tissue processing and automated immunohistochemistry

Whole tissue sections were used for immunohistochemistry instead of tissue microarrays, which limits the area of sampling. A four-micron-thick section was obtained from each FFPE block and stained with hematoxylin and eosin (H and E), with one block per patient. H and E sections were reviewed by study pathologists to confirm the histological status of each FFPE block. Additional four-micron-thick sections were obtained from each FFPE block and stained with PD-L1 (SP263, Ventana) and PD-1 (NAT 105, Ventana Medical Systems, Tucson, AZ) antibodies. Chromogenic staining (DAB single-plex) was performed on the Ventana Discovery Ultra autostainer (Roche Diagnostics, Indianapolis, IN). FFPE sections were mounted on Superfrost Plus microscope slides (Thermo Fisher Scientific, Waltham, MA) and dried overnight prior to staining. After deparaffinizing the slides, antigen retrieval was performed in TRIS-based CC1 buffer (Ventana) at 95°C-97°C for 4 minutes, followed by incubation with primary antibodies. Enzymatic detection of anti-PD-L1 and anti-PD-1 antibodies was performed with a secondary anti-rabbit IgG (Anti-Rabbit HQ RUO Ventana) and anti-mouse IgG (Anti-Mouse HQ RUO Ventana) antibody, respectively. The secondary IgG antibodies were conjugated to HQ, a hapten-labeled conjugate that was detected by DISCOVERY anti-HQ HRP. Chromogenic detection was performed with Chromomap DAB (Roche) followed by counterstaining with hematoxylin. Tonsil sections were stained with and without the primary antibody, serving as positive and negative controls for each marker. As a negative reagent control, specimens were incubated with a rabbit monoclonal (Rabbit Monoclonal Negative Control, IgG, Ventana, 790-4795) and a mouse monoclonal negative control [Negative control (Monoclonal), Ventana, 760-2014] under the same conditions. Image acquisition was performed on a Zeiss Axio Scan.Z1 whole slide scanner. Images were captured with a Zeiss Plan-Apochromat 20x/0.8NA (WD=0.55mm) M27 objective lens in the brightfield mode with Hitachi HV-F202 camera.

### Pathologic assessment of hematoxylin and eosin, PD-L1 and PD-1 staining

96 stained slides were assessed by two pathologists specializing in lower genital tract pathology. Analyses were blinded to the patient’s HIV status. Each sample was reviewed twice on a standard Olympus BX46 microscope. The percentage of PDL-1 and PD-1 immunoreactive cells in aSCC, aHSIL or benign anal tissue specimens was based on assessment of the either the entire invasive, dysplastic, or benign area of a specimen derived from either biopsies or resections. Areas containing tumor necrosis, poor tissue fixation or crush artifact were not considered for immunohistochemistry staining. Nuclear or granular cytoplasmic staining alone was not considered positive. H and E-stained slides from the same block were used as a reference for scoring PD-L1 and PD-1 positivity.

Quantitative analysis of immunohistochemical positivity was performed according to standard clinical practice as described by Kim et al., using a modified scoring system to quantify PD-L1 and PD-1 expression in anal epithelial cells and ICs (16–18). Anal epithelial cells were considered positive for PD-L1 when they exhibited either partial or complete membrane staining. The Tumor Proportion Score (TPS) was used to assess the percentage of tumor cells positive for PD-L1. TPS was defined as the number of PD-L1-positive tumor cells divided by the total number of tumor cells multiplied by 100. We used the term “PD-L1 Epithelial Staining Score (PD-L1 ESS)” to quantify PD-L1 staining in anal epithelial cells of benign and HSIL lesions. We described PD-1 staining of anal epithelial cells using the term “PD-1 Epithelial Staining Score (PD-1 ESS)”. TPS, PD-L1 ESS and PD-1 ESS were scored as follows: ≥5%, 5-9%, 10%, 20%, 30%, 40%, 50%, 60%, 70%, 80%, 90%, 100%, with the cut-off being <5%. For values of 5-9% range the lower value 5% was used for analysis (19). TPS was reported as a dichotomous variable (positive vs. negative) and quartiles, expressed as percentage scores. Staining distribution was determined to be focal, patchy or diffuse. Focal distribution was defined as staining in less than 10% of cells in a single or few locations. Patchy stain distribution was defined as staining present in greater than 10% of cells in multiple locations. Diffuse stain distribution was defined as staining present involving a large area/majority of the cells in a continuous pattern of staining.

The total immune cell infiltrate or “Total IC” was determined by H and E analysis and was defined as the number of ICs, including lymphocytes, macrophages and histiocytes present in the stroma and within the epithelium of benign specimens, or pre-cancerous areas of aHSIL specimens, or within tumor nodules of aSCC specimens. In tumor nodules, these were referred to as tumor-infiltrating lymphocytes (TIL). The Total IC was scored as a percentage: <1%, 1%, 5-9%, 10%, 20%, 30%, 40%, 50%, 60%, 70%, 80%, 90%, 100%. For calculation purposes, anything less than 1% was converted to 0.05. While for values in the 5-9% range, the lower value was reported as determined by the pathologist (20).

PD-L1 or PD-1 positivity of ICs was determined by membrane staining, either complete or incomplete, as well as cytoplasmic or punctate staining. ICs were also scored depending on their location either the stroma or epithelium. The percentage of stromal IC or epithelial IC was scored as <5%, 5-9%, 10%, 20%, 30%, 40%, 50%, 60%, 70%, 80%, 90%, 100% with values <5% scored as zero. The total percentage of PD-L1 or PD-1-positive ICs, i.e., Overall-PD-L1 IC or Overall-PD-1 IC, irrespective of location, was defined as the percentage of PD-L1-positive or PD-1-positive cells as a percentage of Total IC. The scoring was as follows <5%, 5-9%, 10%, 20%, 30%, 40%, 50%, 60%, 70%, 80%, 90%, 100% with the cut-off being <5%. For values in the 5-9% range, the lower value was reported as determined by the pathologist.

We also evaluated PD-L1 expression in aSCC using combined positive score (CPS). CPS was defined as the number of PD-L1 staining tumor cells and mononuclear inflammatory cells within tumor nests and/or adjacent supporting stroma divided by the total number of tumor cells X 100. For CPS score the cut-off of ≥1 was considered positive with any score below 1 considered to be zero. CPS was calculated in a range from 0-100. For benign and HSIL samples, we coined the term “Aggregate PD-L1 score (APS)”, defined as PD-L1 staining of both anal squamous epithelial cells and ICs.

The TME was classified into four types according to the criteria established by Teng et al.(13). This classification considered PD-L1 staining in both tumor cells and ICs. To further support the classification, studies were performed to assess the relationship between PD-L1 expression on tumors and ICs with Total IC and PD-1 expression in ICs.

### RNA In situ hybridization

RNAscope (Advanced Cell Diagnostics, ACD Bio) in situ hybridization was performed on four μm thick FFPE sections obtained from aSCC specimens from three patients. RNAscope Multiplex Fluorescent Reagent Kit (ACD Bio, Cat #323100) was utilized. Samples were selected based on the expression of PD-L1, indicating adaptive immune resistance phenotype. This included samples showing high and low PD-L1 expression on ICs alone, as well as in tumor cells or both. Tissues were pre-treated to improve target recovery as recommended by the manufacturer. Probes for human IFN-γ, PD-L1, PD-1 and CD68 mRNA (ACDBio, Cat #310501, 600861-C2, 602021-C3, 560591-C4) were incubated at 1:50 dilution for 2h at 40°C. The probes were then hybridized with Opal 7-Color Manual IHC Kit (PerkinElmer, Cat #NEL811001KT), for detection of IFN-γ, PD-L1, PD-1 and CD68 using Opal 620, Opal 540, Opal 650 and Opal 690 and respectively at 1:700 dilution.

### Immunofluorescence

RNAscope-stained samples were then counterstained with CD3 (Abcam, Cat #ab1669) at 1:100 dilution. Targets were detected using Alexa Fluor 555-conjugated goat anti-rabbit IgG secondary antibody (Southern Biotech, Cat #4050-32) at 1:100. Tissues were counterstained with DAPI and mounted using prolong gold antifade mountant (Invitrogen, Cat #P36930). Slides were imaged at 63X magnification using a Zeiss Axio Scan.Z1 (Zeiss Group). Multiple ROIs from each specimen slide were selected using serial PD-L1 stained sections to guide imaging.

### HPV genotyping

To prepare DNA from the FFPE tissue, an additional 15-micron-thick section was cut from blocks using a sterile technique to avoid cross-contamination. The protocol used for HPV genotyping has been described previously (2). Briefly, the scroll section was dissolved in CitriSolv™ (Decon Labs, Inc., King of Prussia, PA, USA), and the DNA was prepared using the RecoverAll™ Total Nucleic Acid Isolation Kit for FFPE (Thermo Fisher Scientific, Austin, TX, USA). PCR was performed using a modified pool of MY09/MY11 consensus HPV L1 primers as well as primers for amplification of the human beta-globin gene as an indicator of specimen adequacy. After 40 amplification cycles, specimens were probed with a biotin-labeled HPV L1 consensus probe mixture. A separate membrane was probed with a biotin-labeled probe to the human beta-globin gene. Specimens were also typed by hybridizing to 39 different HPV probes consisting of single and multiple HPV types: 16, 18, 31, 33,35, 39,45, 51, 52, 56, 58, 59, 66, 26/69, 30, 34, 53, 67, 68, 70, 73, 82, 85, 97, 6, 11, 54, 61, 62, 32/42, 71, 72, 81, 83, 84, 86/87, 90/106, 102/89, Mix 1 (7, 13, 40, 43, 44, 55, 74, and 91). Specimens negative for beta-globin gene amplification were excluded from analysis. Specimens in which no HPV DNA was detected were assigned as “No HPV”. Specimens in which the HPV genotype could not be determined were assigned as “Type Unknown”. High Risk (HR) HPV genotypes included HPV 16 and all other HR HPV genotypes (18, 31, 33 35, 39, 45, 51, 52, 56, 58, 59, 66, 26, 69, 30, 34, 53, 67, 68, 70, 73, 82, 85, 97). HPV types other than HPV 16 were assigned as “Non-16 HR HPV”.

### Statistical analysis

Descriptive statistics were used to summarize the data. We assessed some variables using the dichotomous version as well as via quartiles using unbiased cut-offs to explore potential non-linear patterns to assess the best fit to the data. CPS and APS were both reported using the cut-off by dichotomizing them as positive (APS≥5%, TPS ≥5%, and CPS ≥1%) or negative (APS < 5%, TPS < 5%, CPS < 1%) and as quartiles.

We used the Kruskal Wallis test when comparing multiple groups, otherwise we used the Mann-Whitney test for two-group comparisons of numeric outcomes. Spearman correlations were also evaluated for numeric variables. We used chi-square, or Fisher’s exact test as appropriate to compare categorical variables. If variables were not statistically significant (2-sided P<0.05), we did not consider them in further analyses.

The Kaplan-Meier survival curves were generated for survival outcomes and groups were stratified and compared using the log-rank statistic. Hazard ratios (HR) and 95% confidence intervals (CI) were produced from Cox proportional hazards models for survival outcomes. Proportional hazards (PH) was checked using SAS 9.4 phreg using zph option, and all variables met the PH assumption. Due to the low number of events, we did not adjust the Cox model for potential confounders, thus limiting our analyses to univariable analyses. Given the exploratory nature of this study, multiple comparison adjustments were not considered. 2-sided p-values <0.05 were considered statistically significant. Power calculations were not conducted beforehand, and the cohort sampled was based on the availability of tissue specimens. All analyses were generated using SAS v. 9.4.

## Data Availability

The data generated in this study are available upon request from the corresponding author.

## Results

### Demographic and clinical characteristics of study participants stratified by diagnosis

The demographic and clinical characteristics of study participants stratified by histopathologic diagnosis are shown in **Table 1**. 96 individuals contributed tissues including 33 (34.3%) females and 63 males (65.6%) with the following diagnoses: 22 benign (22.9 %), 22 HSIL (22.9%) and 52 SCC (54.2%). The age range was 25 to 77 years with a median age of 55 years. The age distribution showed notable differences among the diagnostic categories. In the 50-69 age bracket, a higher prevalence of aSCC (73.1%, 38/52) cases was observed. No significant differences were noted across the groups in terms of immune status. Among all the ethnicities the White participants comprised the largest proportion: 45.5% (10/ 22) of benign cases, 68.2% (15/22) of HSIL cases, 73.1% (38/52) of SCC cases. Most of the biopsies and resection specimens were taken from the anal canal. HPV 16 was the most prominent type, either alone or in combination with other HPV types. The proportion of infection with HPV 16 alone varied between diagnostic groups. Nearly half of HSIL and SCC samples were infected with only HPV 16. Other high-risk types (non-HPV 16) were present, and aHSIL samples contained a higher proportion of non-HPV 16 HR genotypes compared to benign and aSCC. The characteristics of the aSCC population, stratified by HIV status, showed that sex, history of prior treatment (chemoradiation or surgery), and lesion location were statistically significant between the two groups (Supplementary Table S1).

**Table 1:** Demographic and clinical characteristics of study participants stratified by diagnosis.

|  |  |  | **Anal Benign**<br>**(N=22)** | **Anal HSIL**<br>**(N=22)** | **Anal SCC**<br>**(N=52)** | **Fisher's exact,**<br>**P-value**<br>**(Three group**<br>**comparisons)** |
| --- | --- | --- | --- | --- | --- | --- |
| **Sex** | Female |  | 5 (22.7%) | 9 (40.9%) | 19 (36.5%) | 0.39 |
|  | Male |  | 17 (77.3%) | 13 (59.1%) | 33 (63.5%) |  |
| **Age group** | 25-49 |  | 11 (50.0%) | 13 (59.1%) | 9 (17.3%) | 0.0019 |
|  | 50-69 |  | 10 (45.5%) | 7 (31.8%) | 38 (73.1%) |  |
|  | 70+ |  | 1 (4.5%) | 2 (9.1%) | 5 (9.6%) |  |
| **Immune Status** | HIV-negative, non-immunosuppressed |  | 10 (45.5%) | 10 (45.5%) | 23 (45.1%) | 1.00 |
|  | HIV-negative, immunosuppressed |  | 1 (4.5%) | 1 (4.5%) | 2 (3.9%) |  |
|  | People living with HIV |  | 11 (50.0%) | 11 (50.0%) | *26 (51.0%) |  |
| **Race** | Asian |  | 5 (22.7%) | 1 (4.5%) | 0 | 0.044 |
|  | Black |  | 4 (18.2%) | 3 (13.6%) | 6(11.5%) |  |
|  | Hispanic |  | 3 (13.6%) | 2 (9.1%) | 4 (7.7%) |  |
|  | Mixed |  | 0 | 0 | 1 (1.9%) |  |
|  | Unknown |  | 0 | 1 (4.5%) | 3 (5.8%) |  |
|  | White |  | 10 (45.5%) | 15 (68.2%) | 38 (73.1%) |  |
| **\*\*Location of Biopsy/Resection** | Anal |  | 9 (75.0%) | 18 (90.0%) | 39 (75.0%) | <0.0001 |
|  | Perianal |  | 3 (25.0%) | 2 (10.0%) | 13 (25.0%) |  |
| **HPV** | HPV | present | 10 (45.5%) | 22 (100%) | 50 (96.2%) | <0.0001 |
|  |  | absent | 12 (54.5%) | 0 | 2 (3.8%) |  |
|  | HPV 16 only | present | 0 | 11 (50%) | 25 (48.1%) | <0.0001 |
|  |  | absent | 22 (100%) | 11 (50%) | 27 (51.9%) |  |
|  | HPV 16 with one or more other types | present | 1 (4.5%) | 15 (68.2%) | 40 (76.9%) | <0.0001 |
|  |  | absent | 21 (95.5%) | 7 (31.8%) | 12 (23.1%) |  |
|  | <sup>Δ</sup>Non-HPV 16 with one or more other types | present | 1 (4.5%) | 9 (40.9%) | 19 (36.5%) | 0.0069 |
|  |  | absent | 21(95.5%) | 13 (59.1%) | 33 (63.5%) |  |

\*No information: 1 SCC, \*\* No information: 10 benign and 2 HSIL, <sup>Δ</sup>Infection exclusive of HPV 16

### PD-L1 and PD-1 expression in benign anal epithelium, aHSIL and aSCC

Biopsy tissues from benign, HSIL and SCC lesion were stained with anti-PD-L1 (**Figure 1A, C**) and anti-PD-1 antibodies (**Figure 1B, D**). When samples had more than one histopathological condition, the more severe diagnostic category was considered. Benign anal epithelium showed no expression of PD-L1 in both the epithelium and ICs. A distinct pattern of PD-L1 expression was observed in the aSCC sample, with diffuse PD-L1 expression on the tumor cells, whereas no PD-L1 expression was detected in the anal epithelial cells of aHSIL. Most of the PD-L1-expressing ICs were found near the tumors’ invasive margins. Intratumoral PD-L1 ICs were relatively fewer compared to stromal ICs in aSCC. In HSIL, very few PD-L1 ICs were observed in the intraepithelial region, with the majority located in the stroma.

**Figure 1:**
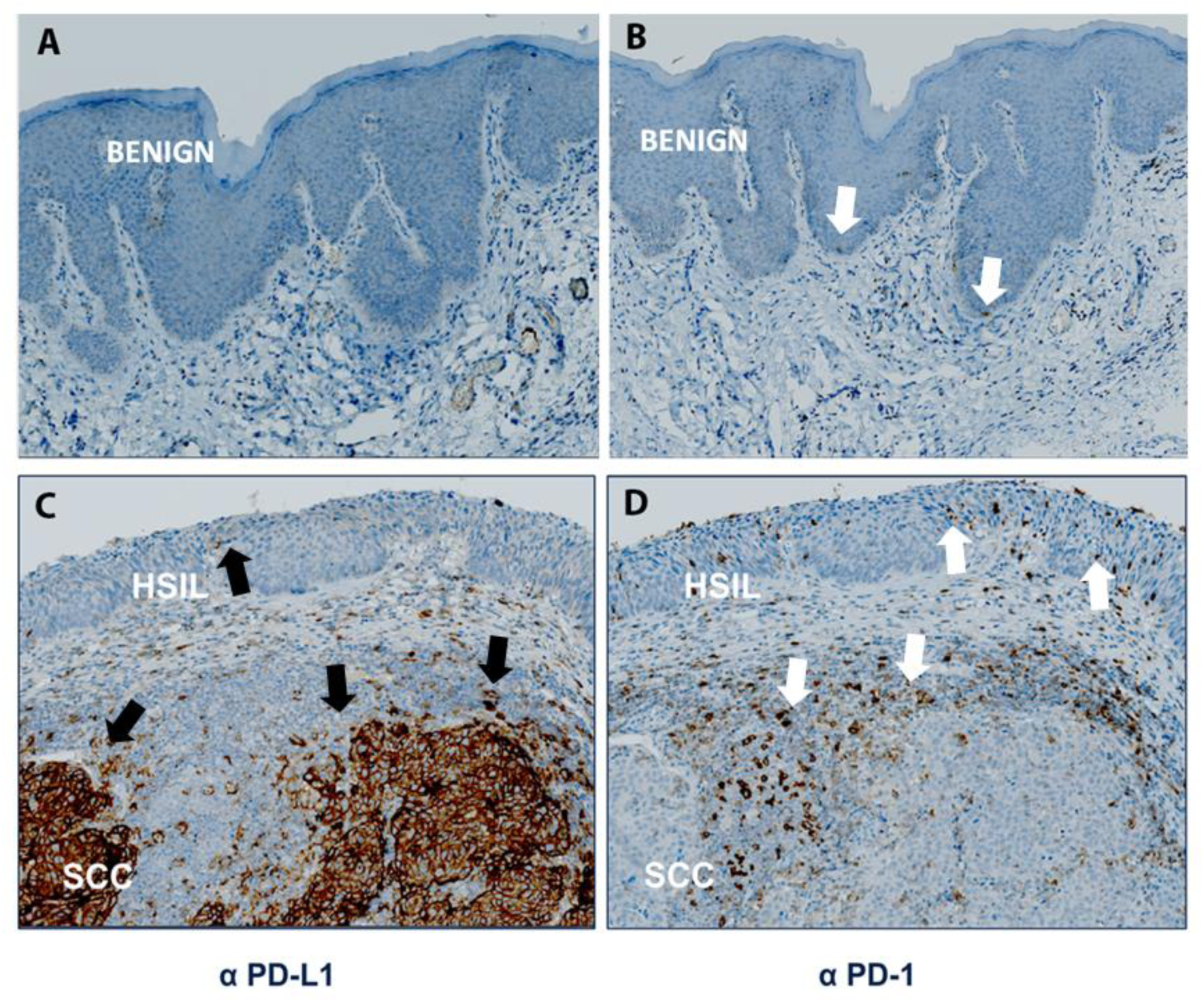
Images of anal benign epithelium, anal HSIL (aHSIL), and anal SCC (aSCC) stained with anti-PD-L1 and anti-PD-1 antibodies. Benign anal epithelial cells are negative for PD-L1, while a few ICs are positive for PD-1 (white arrows) (A, B). Lesion containing both aHSIL and aSCC (C, D). Tumor cells of SCC are PD-L1-positive depicting a distinct PD-L1 expression pattern-diffuse. No PD-L1 expression is observed in epithelial cells in the HSIL region. PD-L1-expressing immune cells (ICs) are mainly located around the tumor-invasive margin and in the stroma (black arrows). PD-1 expression is restricted to ICs and mainly seen around the tumor-invasive margin, correlating with the distribution of PD-L1-positive ICs (white arrows). A high number of PD-1 ICs are observed in the intraepithelial region of aHSIL (white arrows), in comparison to PD-L1 IC (black arrows). [ benign n=22, aHSIL n=22 and aSCC n=52, (20X)].

PD-1 expression was limited to ICs and absent in anal epithelial cells in benign, HSIL and SCC lesions. PD-1-expressing ICs were predominantly situated around the tumor’s invasive margin, similar to the distribution of PD-L1-positive ICs. A substantial number of PD-1-positive ICs were noted in the intraepithelial region of HSIL, but fewer were found in the intraepithelial and stromal region of benign tissues.

### Quantification of PD-L1 and PD-1 positivity in anal epithelial cells and Immune Cells (ICs)

PD-L1 staining in anal epithelial cells was not detected in benign tissues and only 1 of 22 HSIL samples was positive for PD-L1, with 5% of epithelial cells positive for PD-L1. The PD-L1-ESS, TPS APS and CPS are shown in Figure 2. Differences in PD-L1 staining of anal epithelial cells expressed as PD-L1 ESS or TPS (benign=0, HSIL=0, SCC=1) were statistically significant (p=<0.0001) (Figure 2a). The PD-1 Epithelial Staining Score, which indicates PD-1 expression in anal epithelial cells, demonstrated negligible staining across all diagnostic groups. These findings reveal a differential expression pattern of PD-L1 but not PD-1 in epithelial cells across benign, aHSIL, and aSCC lesions.

**Figure 2:**
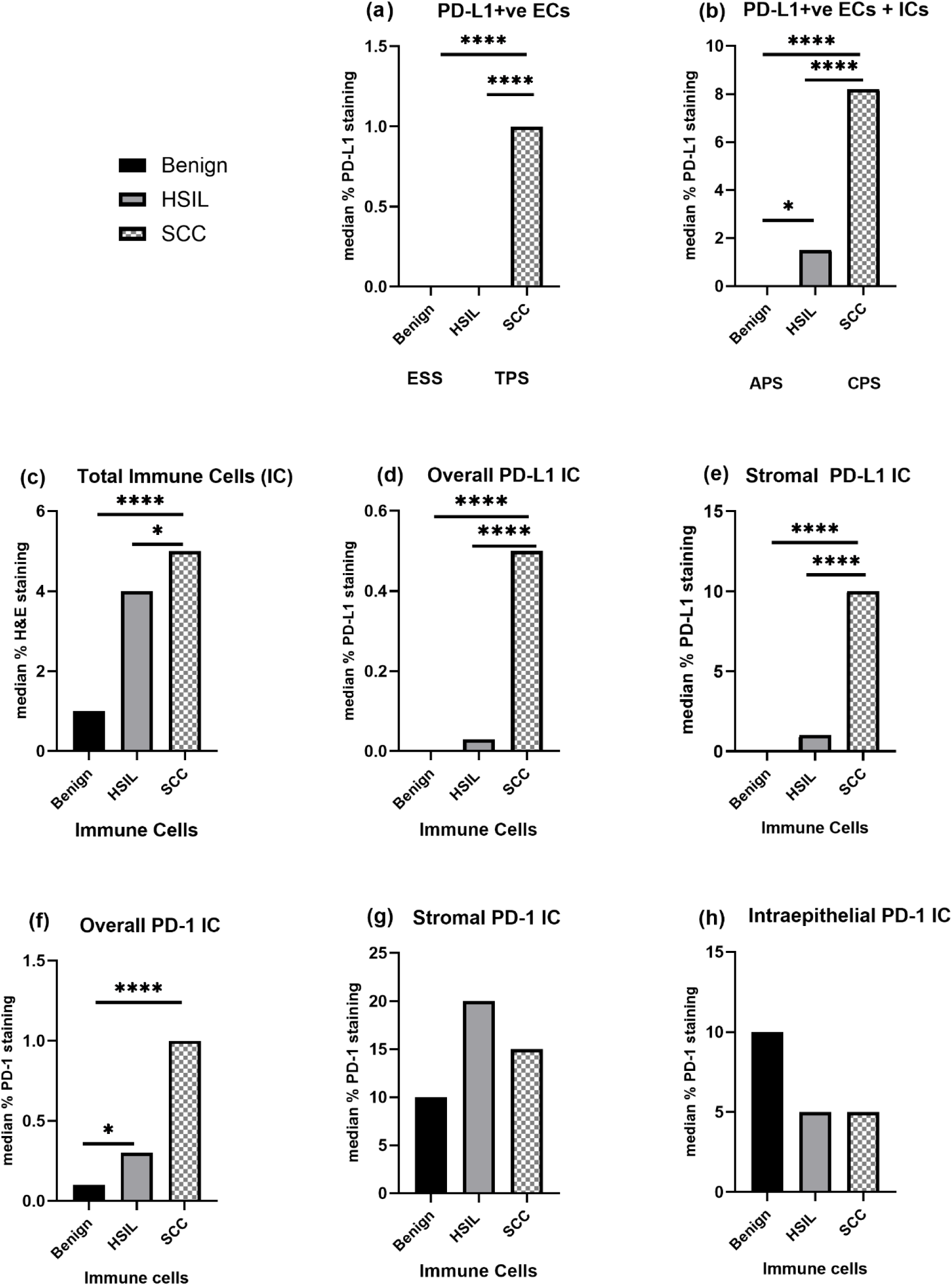
Comparison of hematoxylin and eosin (H&E), PD-L1, and PD-1 expression patterns between histopathological groups: benign n=22, anal HSIL (aHSIL) n=22 and anal SCC (aSCC) n=52 **(a)** PD-L1 Epithelial Staining Score (ESS) of benign and HSIL tissues compared to Tumor Proportion Score (TPS) of SCC. **(b)** Aggregate PD-L1 Score (APS) of benign and HSIL tissues compared to Combined Positive Score (CPS) of SCC. **(c)** Percentage of H&E-positive ICs **(d)** Overall-PD-L1 IC: Percentage of PD-L1-positive IC relative to Total IC. **(e)** Percentage of PD-L1-positive ICs present in the stroma. **(f)** Overall-PD-1 IC: Percentage of PD-1-positive IC relative to Total IC. **(g)** Percentage of PD-1-positive ICs present in the stroma. **(h)** Percentage of PD-1-positive ICs present in the intraepithelial region. In all comparisons, the median percentage has been reported. The median intraepithelial PD-L1-positive IC was zero in all groups. Statistical significance was determined using Mann-Whitney pairwise comparisons. *: p < 0.05, ****: p < 0.0001.

The percentage of immune cell infiltration or ‘Total IC’ increased from benign to HSIL to SCC (p=<0.0001) (**Figure 2c**). PD-L1-positive ICs present in the stroma differed between the three groups (**Figure 2e**), while negligible PD-L1-positive IC was found in the intraepithelial compartment. The Overall-PD-L1 IC, representing the percentage of PD-L1-positive ICs relative to Total IC, increased from benign to HSIL to SCC (**Figure 2d**). Comparison of median PD-1-positive ICs between the intraepithelial and stromal compartments in all three groups did not reach statistical significance (**Figure 2g, h**). However, the Overall-PD-1 IC, which represents PD-1-positive ICs relative to Total IC, was higher in cancers vs. HSIL or benign (p=0.0098) (**Figure 2f**). In all three diagnostic groups, the level of intraepithelial PD-L1 IC and PD-1 IC was lower than that of stromal PD-L1 IC or PD-1 IC. In benign lesions, PD-L1 staining was predominantly absent, exhibiting a median APS of 0. HSIL cases showed minimal PD-L1 staining, with an APS score of 1.5 (lower quartile=0, upper quartile=6.5). In contrast, SCC cases had a high median CPS score of 8.2 and a wider range from 4.65 to 20 (**Figure 2b**).

Between benign and HSIL samples, a significant difference was observed only in APS (p=0.032) and overall-PD-1 IC (p=0.036) (**Figure 2b, f**). Comparison between benign and SCC revealed statistical differences in all categories except for PD-1 IC, located in the intraepithelial and stromal compartments. Differences were observed between HSIL and SCC for Total IC, PD-L1 ESS and PD-L1-expressing ICs and CPS (**Figure 2**). However, for PD-1 IC the differences did not reach statistical significance. Therefore, PD-L1 expression was predominantly found in ICs in three diagnostic stages but was limited to only anal squamous epithelial cells of tumors. In contrast, PD-1 expression was observed exclusively in ICs across all diagnostic stages.

Tumor cells were PD-L1-positive in 53% of SCC cases and displayed various PD-L1 expression patterns: diffuse (4 cases, 7%), focal (5 cases, 9%), patchy (19 cases, 36%). 20 (38%) SCC cases lacked PD-L1 expression on tumor cells. Notably, PD-L1 expression varied between tumors and within the same tumor, appearing either centrally within the tumor (mid-tumor nodule) or at the tumor-stroma interface. In total, 47 tumors (90%) exhibited PD-L1 positivity in ICs. These ICs were located intratumorally, within the stroma, or at the interface between the stroma and tumor. Four tumors lacked PD-L1 expression in both tumor cells and ICs. PD-1-positive ICs were located either intratumorally, within the stroma, or at the tumor-stroma interface.

### PD-L1 expression patterns and TIL distribution in anal SCC reveal distinct tumor microenvironments

Patterns of PD-L1 expression and associated TIL or Total IC were used to characterize the TME. Inducible or adaptive PD-L1 expression was observed in 92% of aSCCs demonstrated by focal or patchy PD-L1 expression on tumor cells or tumor cells at the tumor-stroma interface. PD-1-positive ICs were located adjacent to the PD-L1-positive tumor cells and within tumor nests. PD-L1-positive ICs were found in the stroma, surrounding the tumor margin and a few were found within the tumor nest (**Figure 3A, B**). In these tumors PD-L1 expression on ICs correlated with the degree of Total IC *(r*=0.6, p=0.0014), and PD-1 IC (r=0.7, p=0.0001) (Supplementary Table S2). Additionally, in some tumors, PD-L1 positivity was exclusively observed on ICs located in the stroma adjacent to PD-1-positive ICs. PD-L1 expression on ICs correlated with the degree of Total IC *(r*=0.9, p<0.001), and PD-1 IC (*r*=0.8, p<0.001). Three tumors out of 52 (5.8%) exhibited PD-L1 negativity in both epithelial and ICs despite being positive for Total IC and PD-1-positive ICs. These tumors were designated as having a “tolerance” phenotype (**Figure 3C, D**). In 4 tumors (7.7% of SCC cases), intrinsic induction or constitutive PD-L1 expression was observed. These SCCs displayed a diffuse PD-L1 expression pattern with a TPS score ranging from 60% to 100%. PD-L1 expression on ICs had a low negative correlation with the degree of Total IC (*r*=-0.3, p=0.74) but correlated with PD-1-positive ICs (*r*=0.8, p=0.2), although it did not reach statistical significance (Supplementary Table S2). Notably, these tumors also showed features of adaptive PD-L1 expression, as evidenced by positive Total IC. (**Figure 3E, F**). PD-L1 expression on tumor cells had a weak negative correlation with Total IC or Overall PD-1 IC for SCCs displaying adaptive or mixed PD-L1 expression patterns. However, the correlation did not reach statistical significance. Therefore, distinct patterns of PD-L1 and PD-1 expression in aSCC indicate variations in the TME depending on the immune response.

**Figure 3:**
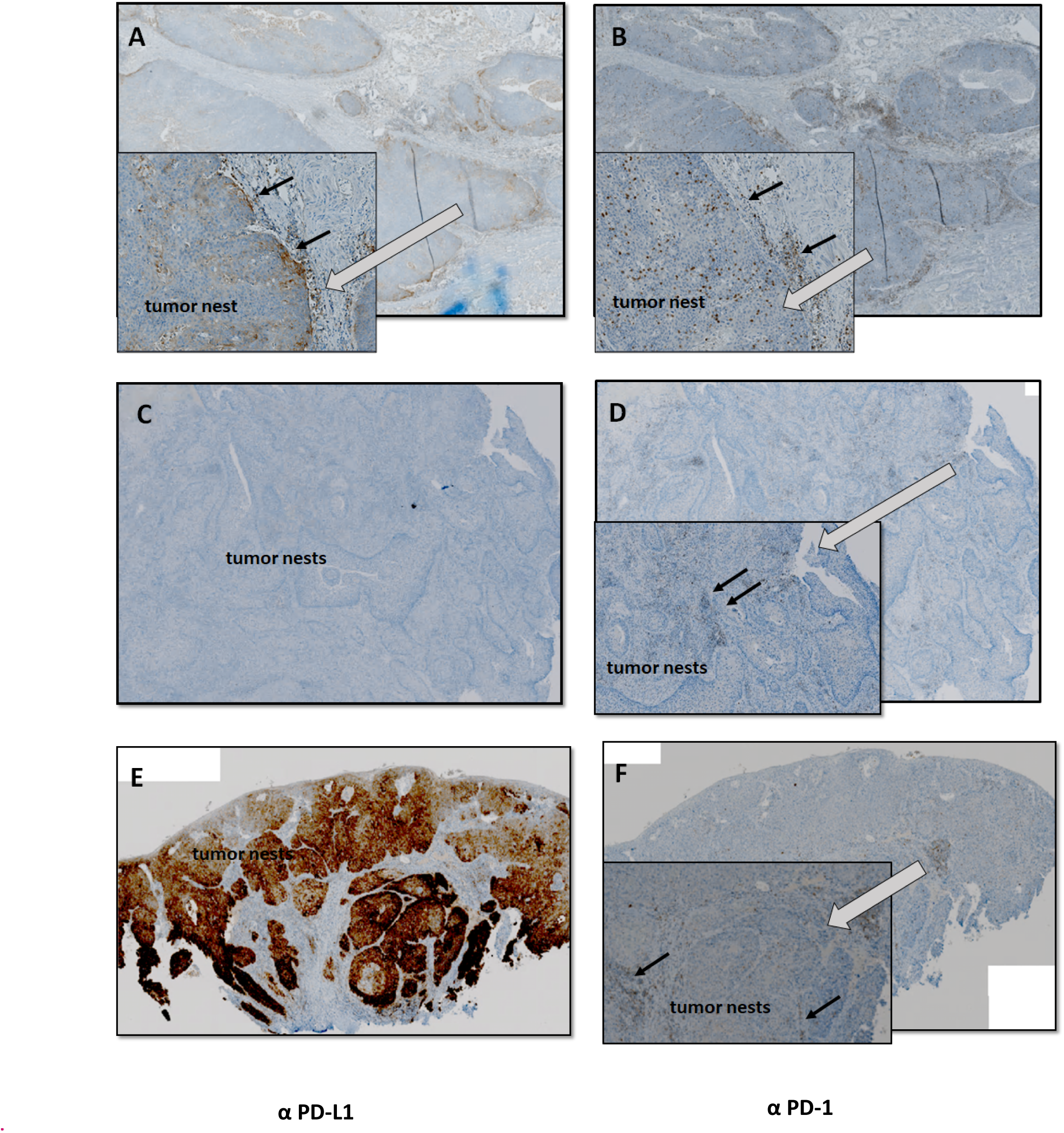
PD-L1 and PD-1 expression patterns in anal squamous cell carcinoma (aSCC) (A) **Adaptive or inducible PD-L1 expression** demonstrated by peripheral and focal PD-L1 expression on tumor cells and PD-L1-positive ICs. PD-L1-positive ICs are found within the tumor and surrounding the tumor. Inset A: Part of a tumor nest enlarged depicting PD-L1-positive tumor cells at the periphery and PD-L1-positive ICs specifically in the stroma (B) PD-1-positive ICs are found in the intratumoral region and in the stroma. Inset B: Enlarged section of a tumor nest showing a high number of intratumoral PD-1-positive ICs and stromal PD-1-positive ICs. (C) **Immunological tolerance** is defined by no PD-L1 expression on tumor cells and ICs. (D) Presence of PD-1-positive ICs. Inset D: Enlarged tumor nests showing a few PD-1-positive immune cells, mostly located in the stroma. (E) **Constitutive PD-L1 expression or intrinsic induction** is demonstrated by diffuse expression of PD-L1 in tumor cells. (F) Presence of PD-1-positive IC. Inset F: Enlarged tumor nests showing low number of PD-1-positive ICs. White arrows point to the enlarged section of the tumor. Black arrows indicate PD-L1-positive tumor cells and ICs (A), PD-1-positive ICs (B, D, F). [aSCC n=52, (20X)].

### Association of TPS and CPS with clinical characteristics and survival outcome of SCC participants

The association between PD-L1 positivity and negativity with clinical characteristics of SCC patients was investigated. Neither TPS nor CPS association with patient characteristics (i.e., sex, age, race, immune status, tumor stage, and tumor grade) reached statistical significance (Supplementary Table S3). The five-year overall survival (OS) rate for this cohort was 74%. While there was no difference in overall survival between PLWH and HIV-negative individuals (log-rank P=0.9678), variations were observed across different tumor stages (log-rank P=0.0007) (Supplementary Figure S1). OS was assessed by PD-L1 positivity in tumor cells and ICs. When TPS was analyzed as either positive or negative or by quartile (0, 1-5, 5-10, >10), higher scores were associated with improved survival but these results did not achieve statistical significance (log rank P=0.65 and log rank P=0.52) (**Figure 4**). However, for those with a positive CPS, higher overall survival and PFS were found compared to those with a negative CPS (log-rank P=<0.0001) and PFS (log rank P=0.0099) (**Figure 5**). Higher scores were also associated with improved survival and PFS when CPS was categorized by quartile (0-4.5, 4.5-8.1, 8.1-20, >20) but these results did not reach statistical significance (log rank P=0.14 and log rank P=0.15).

**Figure 4:**
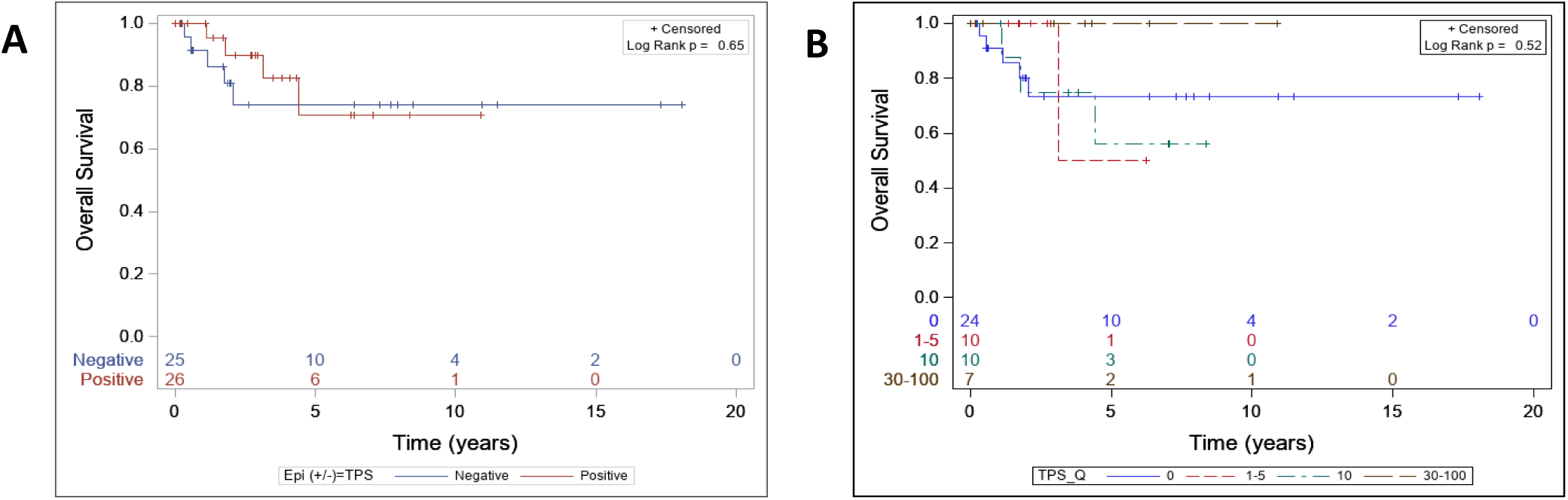
Overall survival (OS) stratified by tumor proportion score (TPS). TPS as a dichotomous variable (positive vs negative) **(A)** and quartiles (0%, 1%-5%, 5%-10%, >10%) **(B).** [aSCC, N=51]

**Figure 5:**
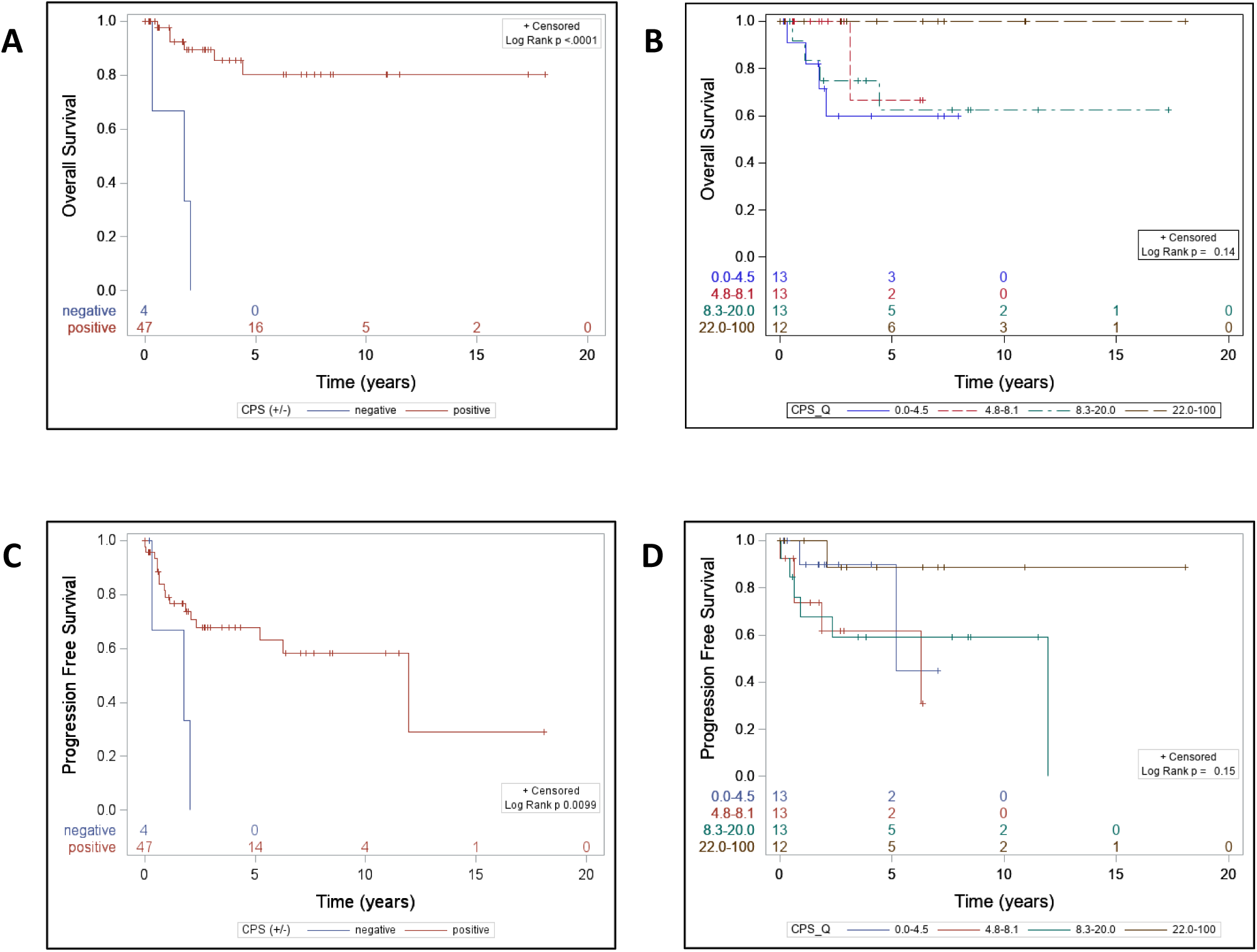
Kaplan Meier analysis stratified by combined positive score (CPS). CPS is categorized into dichotomous variable (positive vs negative) (**A, C)** quartiles (0%-4.5%, 4.5%-8.1%, 8.1%-20%, >20%) **(B,D,)** Overall survival (OS) **(A,B)**; Progression free survival (PFS) **(C,D).** [aSCC, N=51]

### Cox proportional hazard models

CPS was a favorable prognostic marker for aSCC. Negative CPS significantly impacted overall survival compared with positive CPS, with a hazard ratio (HR) of 15.2 (95% CI: 3.3 to 69, p=0.0004; Log-Rank P<0.0001). Negative CPS values were associated with worse PFS (HR: 1.96, 95% CI: 0.57 to 6.8, p=0.28) but these results did not reach statistical significance (Supplementary Table S4b). However, CPS when assessed as a quartile variable, the association with PFS was as follows: if CPS is less than 20, the risk of progression or death is estimated to be 6 to 8 times higher compared to those with CPS >20 (Supplementary Table S4a).

### Interferon-γ expression in anal tumor microenvironment

PD-L1 expression can be either tumor cell-intrinsic or as an adaptive mechanism in response to IFN*-*γ. Because we saw improved overall survival with PD-L1 expression, we hypothesized that PD-L1 expression was driven by an adaptive response reflecting IFN*-*γ production. To test this, we performed in situ hybridization for IFN*-*γ in conjunction with immunofluorescence. Three SCC samples displaying adaptive PD-L1 expression by immunohistochemistry were probed with RNA probes targeting PD-L1, PD-1, CD68, and IFN*-*γ mRNA. T cells were identified with CD3 antibody. No staining was observed with negative control probes (Supplementary Figure S2). A representative anal TME shows PD-L1-positive cells as well as cells co-expressing PD-L1 and CD68. IFN*-*γ was present in the anal TME colocalizing with both CD3+ T cells and within a subset of double-positive CD3+ PD-1-positive T cells. These cells were found in proximity to CD68-positive PD-L1-expressing macrophages and PD-L1-positive cells. These data depict an inflamed TME with macrophages, T cells and IFN*-*γ (**Figure 6**).

**Figure 6:**
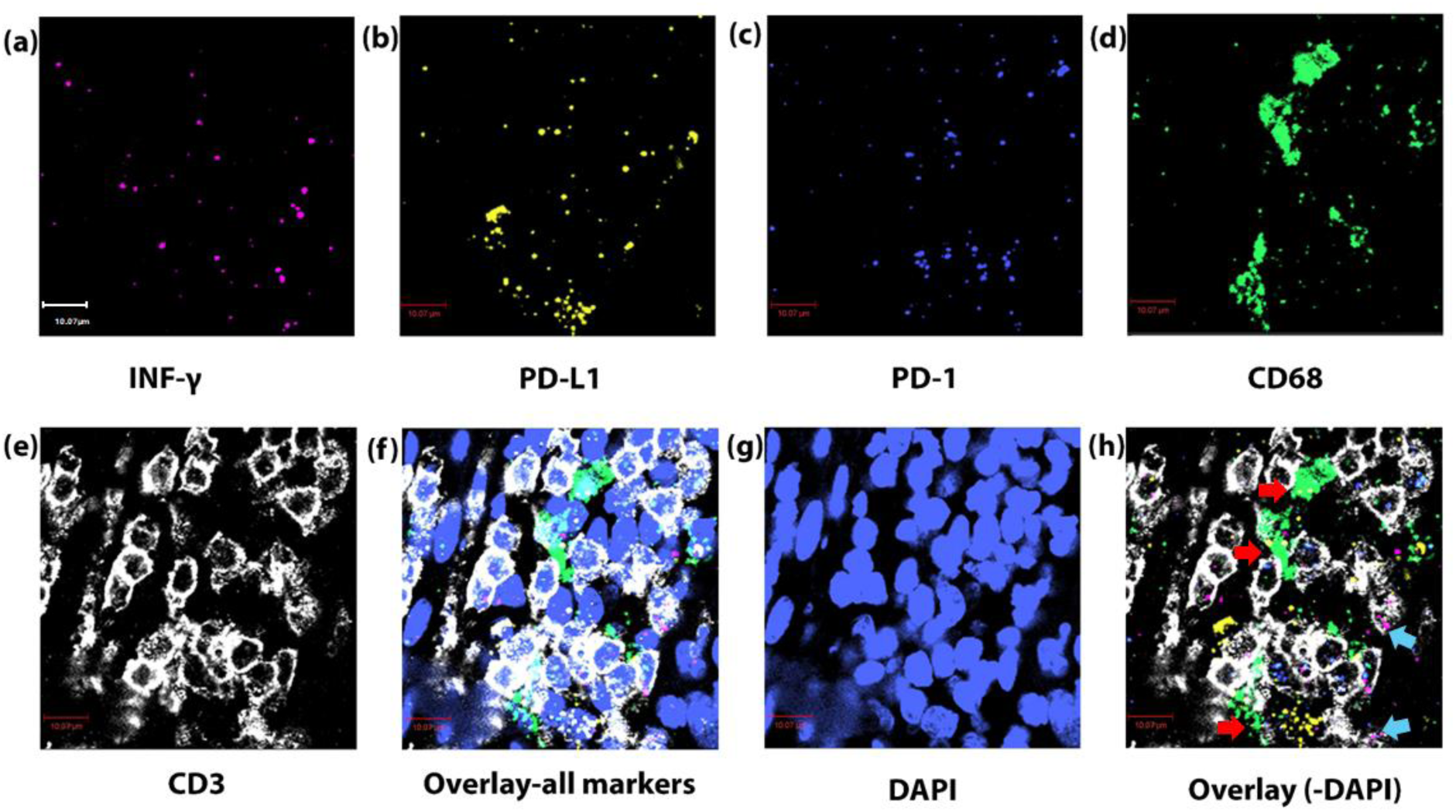
In situ hybridization and multiplex immunofluorescence staining of a representative TME of an anal SCC (aSCC). a) IFN-γ (RNAscope probe, pink), b) PD-L1 (RNAscope probe, yellow), c) PD-1 (RNAscope probe, blue), d) CD68 (RNAscope probe, green), e) CD3+ (immunohistochemistry, white), f) Overlay with DAPI (blue), g) DAPI h) Overlay without DAPI. CD68+ cells that co-express PD-L1 (red arrows), CD3+ cells that co-express PD-1 and IFN-γ (blue arrows), scale=10.07 μm [63X (aSCC, N=3)].

## Discussion

The aim of our study was to examine PD-L1/PD-1 expression patterns in anal tissue (benign, HSIL and cancer), in both PLWH and HIV-negative participants. All samples were evaluated under standardized staining conditions. In contrast, most studies have primarily investigated PD-L1 expression in aSCC and only one study investigated the role of PD-L1 in aHSIL. Unlike these studies that focus solely on PD-L1 expression, we examined both PD-L1 and PD-1, thus offering a more complete understanding of the PD-L1/PD-1 axis in anal disease.

Epithelial PD-L1 positivity was nearly exclusively observed in aSCC, with only one aHSIL sample exhibiting PD-L1 positivity and no benign samples exhibiting PD-L1 expression. In ICs the PD-L1 positivity increased from HSIL to SCC. Histological analysis and subsequent RNA in situ hybridization on SCC samples identified these ICs primarily as macrophages. We found a substantial rise in the APS/CPS score, from 1.5 in HSIL to 8.2 in SCC when comparing HSIL and SCC lesions. This increase was accompanied by elevated PD-L1 positivity in ICs, and significantly, PD-L1 positivity was observed in anal epithelial cells of aSCC. In contrast, PD-1 positivity between HSIL and SCC did not reach statistical significance. These findings suggest that upregulation of PD-L1 and immunomodulation via the PD-L1/PD-1 pathway in tumor cells compared with HSIL may be an important step in transitioning from HSIL to cancer.

Our data contrasts with a prior study in which PD-L1-positive epithelial cells were observed in one (6%) AIN1 sample and 7 (12%) AIN2/3 samples. This disparity in results may stem from differences in antibodies used and variations in the interpretation of staining patterns (21). In cervical HSILs PD-L1 expression has been reported primarily in cervical squamous epithelial cells. In a study by Mezache et al., 95% (20/21) of CIN I-II cases were PD-L1-positive (22). When comparing PD-1 expression in our study to that of Bucau et al., we found that PD-1 was exclusively detected in ICs with an increase in expression observed as the grade of anal disease progressed. Notably, PD-1 positivity was found in 95.5% of HSIL cases, contrasting with Bucau et al., who reported PD-1-positive lymphocytes in only 24 (41%) AIN2/AIN3 samples (21).

A comparison between benign and HSIL samples revealed not only an increase in Total IC (immune infiltrate) between benign and HSIL samples, but also an increase in both the APS score and Overall-PD-1 IC score. These findings suggest that immune suppression may be initiated during the precancerous stage, triggered by activation of the PD-L1/PD-1 pathway (23). Cell-mediated immunity driven by T cells is known to play a major role in regression of HPV-induced pre-malignant lesions (7). Therefore, we hypothesize that the upregulation of PD-L1 in ICs may lead to suppression of PD-1-positive T cell activity. The downregulation of cell-mediated immunity at this critical transition stage may contribute to disease persistence and possibly progression to cancer. However, this hypothesis needs to be further explored through mechanistic experimental studies focusing on PD-L1/PD-1 function in HSIL.

Most aSCC are HPV-driven and have a high frequency of TILs and a substantial inflammatory response. This response is also seen in other HPV-driven cancers, such as head and neck cancer and many other virally driven malignancies (24). In our cohort, 96% of SCCs were positive for HPV, and the tumors were characterized by a high TIL score consisting of PD-1-positive and PD-L1-positive ICs. These tumors can be considered “hot tumors” or “inflamed tumors” due to the significant inflammation within the TME, spurred in part by the presence of E6 and E7 oncoproteins expressed by the tumor cells (25, 26). The Overall-PD-1 IC expression in SCC samples ranged from 0.25% to 1.0% in aSCC. Despite PD-1 being considered a marker of exhaustion, it is also an activation marker and has been shown to be a positive prognostic marker in cancers such as head and neck, ovarian, pancreatic and colorectal (27).

PD-L1 expression as expressed by TPS and CPS was not associated with clinical characteristics of SCC patients. HIV infection did not influence PD-L1 expression in benign, HSIL, or SCC tissue. This finding in SCC is consistent with prior studies, suggesting that HIV status does not impact PD-L1 expression in aSCC, possibly due to patients being on antiretroviral therapy (28, 29).

Recent studies have focused on the association between PD-L1 expression and overall survival in anal cancer patients. Monsurd et al. found PD-L1 status to be an adverse prognostic marker, whereas Wessley et al. observed a correlation with improved median overall survival (17, 28). However, the relationship between PD-L1 and outcomes after chemoradiotherapy (CRT) is debatable. Govindraj, Zhao et al., and Steiniche et al. found PD-L1 expression linked to poorer overall survival post-CRT, while Balermpas, Iseas et al., and Chan et al. reported opposing results (30–35). Chamseddin and Mitra’s studies found no significant correlation between PD-L1 expression and overall survival post-CRT (36, 37). These conflicting reports underscore the complexity of PD-L1’s role as a prognostic marker for treatment response.

In our study, we found a positive association between PD-L1 expression and improved overall survival in our cohort consisting of mostly treatment naïve primary tumors. As there is no universally accepted cut-off for PD-L1 staining, we assessed both traditional dichotomous thresholds (as positive versus negative, TPS ≥5%, and CPS ≥1%) as well as quartiles to assess PD-L1 expression in SCC. This approach offers a potentially more informative and statistically unbiased method of assessment. When analyzing overall survival by TPS whether as a binary or quartile variable, the results did not reach statistical significance but did show that higher levels are associated with improved outcome. This finding highlights the limitations in relying solely on PD-L1 expression on tumor cells. Furthermore, it does not provide a comprehensive understanding of the immune landscape within the TME. To gain a more complete picture, it is essential to consider both PD-L1 expression in tumor cells and ICs (38). This dual assessment allows for a better understanding of the immune response and its potential impact on patient outcomes (39).

In our study CPS emerged as a notable prognostic factor for overall survival. Likewise, CPS also exhibited significant prognostic value for PFS. High CPS scores were associated with better OS and progression-free survival PFS. Similarly, in a study of head and neck squamous cell carcinoma (HNSCC), higher CPS scores were associated with improved survival when treated with pembrolizumab, either alone or in combination with chemotherapy (40).

The adaptive immune resistance phenotype observed in the anal TME may explain the improved overall survival in patients with higher PD-L1 expression. In response to immune pressure, tumors adapt by displaying PD-L1 on tumor cells and ICs to counteract immune recognition and destruction (41). The majority of tumors (92%) within our cohort demonstrated adaptive PD-L1 expression characterized by PD-L1 expression on tumor cells, ICs, or both. Correlative analyses revealed a significant association between PD-L1 expression on ICs and the presence of tumor-infiltrating PD-1-positive ICs. These results further support the notion of TILs driving an adaptive immune response, as observed in melanoma and various epithelial cancers (42). These TILS secrete INF-γ leading to upregulation of PD-L1 in tumor cells and ICs. Consistent with this, we detected interferon within the anal TME, released by CD3+ PD-1-positive T cells. Notably, these cells were found near PD-L1-positive tumor cells and PD-L1-positive macrophages. Additionally, ICs in the TME more frequently exhibited PD-L1 expression, likely as a result of IFN-γ–induced adaptive regulation, and this correlated with higher number of TILs. Although previous studies have detected interferon in the anal TME using RT-PCR, this method lacks spatial resolution compared to the in-situ hybridization technique used in our study (43). The above data highlight that PD-L1 expression may serve as an indirect marker of IC activity within the TME. Its presence indicates the engagement and response of the host immune system against the tumor. Although PD-L1 is essentially an immunosuppressive marker, its expression often suggests the activity of tumor-specific T cells, which recognize cancer antigens and release IFN-γ, leading to induction of PD-L1. This feedback loop contributes to adaptive immune resistance and can be seen as a marker of pre-existing anti-tumor immunity (44).

Based on our findings, we hypothesize that PD-L1 expressed by anal tumor cells and ICs deactivates PD-1-positive T cells, halting their tumor-killing activity and potentially leading to anal carcinogenesis. Thus, PD-L1 expression, in conjunction with TIL presence, serves as a marker of preexisting immunity. Positive PD-L1 expression, as quantified by CPS, demonstrated higher values that correlated with better survival outcomes. In contrast, negative CPS, was associated with poorer survival. It can be hypothesized that these treatment-naïve PD-L1 positive tumors will respond to chemo-radiation, as suggested by other studies (30, 31, 33, 45). High PD-L1 expression may create an immune-modulating environment that enhances CRT effectiveness through immune-mediated tumor cell death. Moreover, PD-L1 positivity could signify a pre-existing anti-tumor immune response that CRT can further activate, potentially improving treatment outcomes (30, 46).

Therapy for aSCC includes chemoradiation, surgery and immunotherapy, mostly in locally advanced and metastatic cases (14, 47). Due to the high levels TILS and PD-L1 expression in the anal TME, anal cancers are prime candidates for PD-L1 immunotherapy. The premise of immunotherapy for anal cancer is based on immunologically active TME of aSCCs. Our results also showcased an active TME in primary early stage aSCC. Specifically, survival curves demonstrated that the presence of PD-L1 positivity is associated with better overall survival and PFS. Considering the promising prognostic role of PD-L1/PD-1 immunotherapy in metastatic tumors, it is reasonable to suggest potential benefits for primary tumors as well. However, clinical trials are needed to validate this hypothesis, along with the results from the ECOG-ACRIN 2165, AMC-110 and RADIANCE trials that focus on early-stage aSCC (15).

Our study has some limitations. The retrospective design might have introduced selection biases. A larger cohort would have enabled us to conduct multivariable analyses that allows us to adjust for potential confounders. For this study patients were not treated with immunotherapy and therefore it is essential to validate the observed effects on OS in our study with prospective clinical trials that include immunotherapy. This study also highlighted that the PD-L1/PD-1 axis is active even during the precancerous stage. To investigate whether PD-L1/PD-1 expression influences HSIL progression, comparative analyses between lesions that progress and those that regress are necessary. These efforts will significantly contribute to our understanding of the PD-L1/PD-1 axis within the precancerous microenvironment, shedding light on its implications for progression from HSIL to cancer and for immunotherapeutic strategies in aHSIL. It should be noted that the TME classification used for this study is primarily based on histopathological findings, which is limited in resolution and scope. To gain a more accurate understanding of the dynamic and heterogeneous TME, the application of new analytical methods is essential.

In summary, this study is the first detailed examination of the PD-L1/PD-1 axis in all three histopathological groups (benign anal tissues, aHSIL, and aSCC) along a continuum of disease progression. We demonstrated that PD-L1 expression is restricted to ICs in the precancerous stage and hypothesized that it could be a contributing factor in the induction of anal carcinogenesis. Our data showed that the expression of PD-L1 in anal epithelial cells is only observed in tumor cells of aSCC and upregulation of PD-L1 expression on epithelial cells may be a key step in progression from HSIL to cancer. In the anal TME, PD-L1 expression is higher in ICs than in tumor cells and likely induced by adaptive regulation. We found that understanding the biological mechanisms driving PD-L1 expression, along with its functional significance, requires considering it in the context of the PD-1-positive immune infiltrate. We observed a positive association between PD-L1 expression and improved overall survival. The study also establishes the importance of CPS as a potential prognostic marker for aSCC, regardless of treatment with immune checkpoint blockers.

## Supporting information

Supplementary Data Anal PD-L1 and PD-1

## References

1. Wang C-CJ, Sparano J, Palefsky JM. Human Immunodeficiency Virus/AIDS, Human Papillomavirus, and Anal Cancer. Surgical oncology clinics of North America. 2017;26(1):17–31.

2. Chowdhury S, Darragh TM, Berry-Lawhorn JM, Isaguliants MG, Vonsky MS, Hilton JF, et al. HPV Type Distribution in Benign, High-Grade Squamous Intraepithelial Lesions and Squamous Cell Cancers of the Anus by HIV Status. Cancers. 2023;15(3):660.

3. Fléjou JF. An update on anal neoplasia. Histopathology. 2015;66(1):147–60.

4. Nunes RAL, Morale MG, Silva GÁ F, Villa LL, Termini L. Innate immunity and HPV: friends or foes. Clinics (Sao Paulo). 2018;73(suppl 1):e549s.

5. Shi HJ, Song H, Zhao QY, Tao CX, Liu M, Zhu QQ. Efficacy and safety of combined high-dose interferon and red light therapy for the treatment of human papillomavirus and associated vaginitis and cervicitis: A prospective and randomized clinical study. Medicine (Baltimore). 2018;97(37):e12398.

6. Litwin TR, Irvin SR, Chornock RL, Sahasrabuddhe VV, Stanley M, Wentzensen N. Infiltrating T-cell markers in cervical carcinogenesis: a systematic review and meta-analysis. British Journal of Cancer. 2021;124(4):831–41.

7. Wakabayashi R, Nakahama Y, Nguyen V, Espinoza JL. The Host-Microbe Interplay in Human Papillomavirus-Induced Carcinogenesis. Microorganisms. 2019;7(7).

8. Greten FR, Grivennikov SI. Inflammation and Cancer: Triggers, Mechanisms, and Consequences. Immunity. 2019;51(1):27–41.

9. Zhitkevich A, Bayurova E, Avdoshina D, Zakirova N, Frolova G, Chowdhury S, et al. HIV-1 Reverse Transcriptase Expression in HPV16-Infected Epidermoid Carcinoma Cells Alters E6 Expression and Cellular Metabolism, and Induces a Hybrid Epithelial/Mesenchymal Cell Phenotype. Viruses. 2024;16(2):193.

10. Zicari S, Sessa L, Cotugno N, Ruggiero A, Morrocchi E, Concato C, et al. Immune Activation, Inflammation, and Non-AIDS Co-Morbidities in HIV-Infected Patients under Long-Term ART. Viruses. 2019;11(3).

11. Shamseddine AA, Burman B, Lee NY, Zamarin D, Riaz N. Tumor Immunity and Immunotherapy for HPV-Related Cancers. Cancer Discovery. 2021;11(8):1896–912.

12. Ribas A, Hu-Lieskovan S. What does PD-L1 positive or negative mean? J Exp Med. 2016;213(13):2835–40.

13. Teng MW, Ngiow SF, Ribas A, Smyth MJ. Classifying Cancers Based on T-cell Infiltration and PD-L1. Cancer Res. 2015;75(11):2139–45.

14. Ciardiello D, Guerrera LP, Maiorano BA, Parente P, Latiano TP, Di Maio M, et al. Immunotherapy in advanced anal cancer: Is the beginning of a new era? Cancer Treatment Reviews. 2022;105:102373.

15. Dhawan N, Afzal MZ, Amin M. Immunotherapy in Anal Cancer. Curr Oncol. 2023;30(5):4538–50.

16. Czogalla B, Pham D, Trillsch F, Rottmann M, Gallwas J, Burges A, et al. PD-L1 expression and survival in p16-negative and -positive squamous cell carcinomas of the vulva. J Cancer Res Clin Oncol. 2020;146(3):569–77.

17. Wessely A, Heppt MV, Kammerbauer C, Steeb T, Kirchner T, Flaig MJ, et al. Evaluation of PD-L1 Expression and HPV Genotyping in Anal Squamous Cell Carcinoma. Cancers (Basel). 2020;12(9).

18. Kim SW, Roh J, Park CS. Immunohistochemistry for Pathologists: Protocols, Pitfalls, and Tips. J Pathol Transl Med. 2016;50(6):411–8.

19. Reid MD, Bagci P, Ohike N, Saka B, Erbarut Seven I, Dursun N, et al. Calculation of the Ki67 index in pancreatic neuroendocrine tumors: a comparative analysis of four counting methodologies. Modern Pathology. 2015;28(5):686–94.

20. Brouckaert O, Paridaens R, Floris G, Rakha E, Osborne K, Neven P. A critical review why assessment of steroid hormone receptors in breast cancer should be quantitative. Annals of Oncology. 2013;24(1):47–53.

21. Bucau M, Gault N, Sritharan N, Valette E, Charpentier C, Walker F, et al. PD-1/PD-L1 expression in anal squamous intraepithelial lesions. Oncotarget. 2020;11(39):3582–9.

22. Mezache L, Paniccia B, Nyinawabera A, Nuovo GJ. Enhanced expression of PD L1 in cervical intraepithelial neoplasia and cervical cancers. Mod Pathol. 2015;28(12):1594–602.

23. Greeshma LR, Joseph AP, Sivakumar TT, Raghavan Pillai V, Vijayakumar G. Correlation of PD-1 and PD-L1 expression in oral leukoplakia and oral squamous cell carcinoma: an immunohistochemical study. Scientific Reports. 2023;13(1):21698.

24. Ott PA, Piha-Paul SA, Munster P, Pishvaian MJ, van Brummelen EMJ, Cohen RB, et al. Safety and antitumor activity of the anti-PD-1 antibody pembrolizumab in patients with recurrent carcinoma of the anal canal. Annals of oncology : official journal of the European Society for Medical Oncology. 2017;28(5):1036–41.

25. Selimagic A, Dozic A, Husic-Selimovic A, Tucakovic N, Cehajic A, Subo A, et al. The Role of Inflammation in Anal Cancer. Diseases. 2022;10(2).

26. Wang L, Geng H, Liu Y, Liu L, Chen Y, Wu F, et al. Hot and cold tumors: Immunological features and the therapeutic strategies. MedComm (2020). 2023;4(5):e343.

27. Simon S, Labarriere N. PD-1 expression on tumor-specific T cells: Friend or foe for immunotherapy? Oncoimmunology. 2017;7(1):e1364828.

28. Monsrud AL, Avadhani V, Mosunjac MB, Flowers L, Krishnamurti U. Programmed Death Ligand-1 Expression Is Associated With Poorer Survival in Anal Squamous Cell Carcinoma. Arch Pathol Lab Med. 2021.

29. Yanik EL, Kaunitz GJ, Cottrell TR, Succaria F, McMiller TL, Ascierto ML, et al. Association of HIV Status With Local Immune Response to Anal Squamous Cell Carcinoma: Implications for Immunotherapy. JAMA Oncol. 2017;3(7):974–8.

30. Balermpas P, Martin D, Wieland U, Rave-Frank M, Strebhardt K, Rodel C, et al. Human papilloma virus load and PD-1/PD-L1, CD8(+) and FOXP3 in anal cancer patients treated with chemoradiotherapy: Rationale for immunotherapy. Oncoimmunology. 2017;6(3):e1288331.

31. Chan AM, Roldan Urgoiti G, Jiang W, Lee S, Kornaga E, Mathen P, et al. The prognostic impact of PD-L1 and CD8 expression in anal cancer patients treated with chemoradiotherapy. Frontiers in Oncology. 2022;12.

32. Govindarajan R, Gujja S, Siegel ER, Batra A, Saeed A, Lai K, et al. Programmed Cell Death-Ligand 1 (PD-L1) Expression in Anal Cancer. Am J Clin Oncol. 2018;41(7):638–42.

33. Iseas S, Golubicki M, Robbio J, Ruiz G, Guerra F, Mariani J, et al. A clinical and molecular portrait of non-metastatic anal squamous cell carcinoma. Transl Oncol. 2021;14(6):101084.

34. Steiniche T, Ladekarl M, Georgsen JB, Andreasen S, Busch-Sørensen M, Zhou W, et al. Association of programmed death ligand 1 expression with prognosis among patients with ten uncommon advanced cancers. Future Sci OA. 2020;6(8):Fso616.

35. Zhao YJ, Sun WP, Peng JH, Deng YX, Fang YJ, Huang J, et al. Programmed death-ligand 1 expression correlates with diminished CD8+ T cell infiltration and predicts poor prognosis in anal squamous cell carcinoma patients. Cancer Manag Res. 2018;10:1–11.

36. Chamseddin BH, Lee EE, Kim J, Zhan X, Yang R, Murphy KM, et al. Assessment of circularized E7 RNA, GLUT1, and PD-L1 in anal squamous cell carcinoma. Oncotarget. 2019;10(57):5958–69.

37. Mitra D, Horick NK, Brackett DG, Mouw KW, Hornick JL, Ferrone S, et al. High IDO1 Expression Is Associated with Poor Outcome in Patients with Anal Cancer Treated with Definitive Chemoradiotherapy. Oncologist. 2019;24(6):e275–e83.

38. Kim HR, Ha SJ, Hong MH, Heo SJ, Koh YW, Choi EC, et al. PD-L1 expression on immune cells, but not on tumor cells, is a favorable prognostic factor for head and neck cancer patients. Sci Rep. 2016;6:36956.

39. Ulas EB, Hashemi SMS, Houda I, Kaynak A, Veltman JD, Fransen MF, et al. Predictive Value of Combined Positive Score and Tumor Proportion Score for Immunotherapy Response in Advanced NSCLC. JTO Clin Res Rep. 2023;4(9):100532.

40. Burtness B, Harrington KJ, Greil R, Soulières D, Tahara M, de Castro G, Jr., et al. Pembrolizumab alone or with chemotherapy versus cetuximab with chemotherapy for recurrent or metastatic squamous cell carcinoma of the head and neck (KEYNOTE-048): a randomised, open-label, phase 3 study. Lancet. 2019;394(10212):1915–28.

41. Pardoll DM. The blockade of immune checkpoints in cancer immunotherapy. Nat Rev Cancer. 2012;12(4):252–64.

42. Presti D, Dall’Olio FG, Besse B, Ribeiro JM, Di Meglio A, Soldato D. Tumor infiltrating lymphocytes (TILs) as a predictive biomarker of response to checkpoint blockers in solid tumors: A systematic review. Critical Reviews in Oncology/Hematology. 2022;177:103773.

43. Kowanetz M, Zou W, Gettinger SN, Koeppen H, Kockx M, Schmid P, et al. Differential regulation of PD-L1 expression by immune and tumor cells in NSCLC and the response to treatment with atezolizumab (anti-PD-L1). Proc Natl Acad Sci U S A. 2018;115(43):E10119–e26.

44. Kalbasi A, Ribas A. Tumour-intrinsic resistance to immune checkpoint blockade. Nat Rev Immunol. 2020;20(1):25–39.

45. Gennen K, Käsmann L, Taugner J, Eze C, Karin M, Roengvoraphoj O, et al. Prognostic value of PD-L1 expression on tumor cells combined with CD8+ TIL density in patients with locally advanced non-small cell lung cancer treated with concurrent chemoradiotherapy. Radiat Oncol. 2020;15(1):5.

46. Martin D, Rodel F, Balermpas P, Rodel C, Fokas E. The immune microenvironment and HPV in anal cancer: Rationale to complement chemoradiation with immunotherapy. Biochim Biophys Acta Rev Cancer. 2017;1868(1):221–30.

47. Bian JJ, Almhanna K. Anal cancer and immunotherapy-are we there yet? Transl Gastroenterol Hepatol. 2019;4:57.

