## Supplementary Data Anal PD-L1 and PD-1 for "Programmed death ligand-1 and Programmed cell death protein-1 expression across the anal disease continuum and association with improved survival in anal cancer"

#### Table S1: Characteristics of SCC population: stratified by HIV status

| | | HIV status | | |
|---|---|---|---|---|
| | | •Negative (25, 48%) | PLWH (27, 52.0%) | Fisher's Exact P Value |
| **Sex** | Female | 15 (60.0%) | 4 ( 14.8%) | 0.0013 |
| | Male | 10 (40.0%) | 23 (85.2%) | |
| **Age** | 25-49 | 2 ( 8.0%) | 7 (25.9%) | 0.23 |
| | 50-69 | 20 (80.0%) | 18 (66.7%) | |
| | 70+ | 3 (12.0%) | 2 ( 7.4%) | |
| **Location of lesion** | Anal | 22 (88.0%) | 17 (63.0%) | 0.055 |
| | Peri-anal | 3 (12.0%) | 10 (37.0%) | |
| **Race** | Black | 1 ( 4.0%) | 5 (18.5%) | 0.44 |
| | Hispanic | 2 ( 8.0%) | 2 ( 7.4%) | |
| | Mixed | 1 ( 4.0%) | 0 | |
| | Unknown | 2 ( 8.0%) | 1 ( 3.7%) | |
| | White | 19 (76.0%) | 19 (70.4%) | |
| **Prior treatment°** | No | 20 (80.0%) | **26 (96.3%) | 0.020 |
| | Yes | 5 (20.0%) | 0 | |
| **Grade** | Poor | 7 (28.0%) | 4 (14.8%) | 0.40 |
| | Moderate | 11 (44.0%) | 11 (40.7%) | |
| | Well | 7 (28.0%) | 12 (44.4%) | |
| **Stage** | I | 7 (28.0%) | 10 ( 37.0%) | 0.88 |
| | II | 11 (44.0%) | 11 (40.7%) | |
| | III,IV | $^{\Delta}$ 5 (20.0%) | $^{\Delta\Delta}$ 5 (18.5%) | |
| **$CD4^+$ Counts** | < 200 | NA | 5 ( 20.0%) | |
| | 200-500 | | 12 ( 48.0%) | |
| | > 500 | | 8 ( 32.0%) | |
| **HIV Viral load** | ND (undetectable) | NA | 16 (64.0%) | |
| | >20 and < 1000/ml | | 7 (28.0%) | |
| | >1000 | | •• 2 ( 8.0%) | |
| **Antiretroviral Therapy** | No | NA | ~3 ( 12.0%) | |
| | Yes | | 22 ( 88.0%) | |

• 4 patients immunosuppressed; °Prior treatment includes-chemoradiation or surgery, Missing information: **Treatment Prior-1 Patient, $^{\Delta}$Stage- 2 patients HIV-negative, $^{\Delta\Delta}$Stage- 1 patient HIV Positive, •• HIV viral load-2 patients, ~ Antiretroviral therapy-1 patient

### Table S2: Spearman Correlation of PD-L1 expression in tumor cells and immune cells (IC) with Total IC/TIL and PD-1-positive IC

**Total SCC** (N=52)

| | Tumor PD-L1 expression | | Immune cell PD-L1 expression | |
|---|---|---|---|---|
| | Correlation Co-efficient | P Value | Correlation Co-efficient | P Value |
| Total IC | 0.2 | 0.18 | 0.5 | 0.0002 |
| Overall PD-1 IC | 0.2 | 0.28 | 0.6 | <.0001 |

**Adaptive PD-L1 expression**

Focal and Patchy (N=24)

| | Tumor PD-L1 expression | | Immune cell PD-L1 expression | |
|---|---|---|---|---|
| Immune cells | Correlation Co-efficient | P Value | Correlation Co-efficient | P Value |
| Total IC | -0.3 | 0.18 | 0.6 | 0.0014 |
| Overall PD-11C | -0.14 | 0.51 | 0.7 | 0.0001 |

Non-Epithelial PD-L1 expression (N=20)

| | Tumor PD-L1 expression | | Immune cell PD-L1 expression | |
|---|---|---|---|---|
| Immune cells | Correlation Co-efficient | P Value | Correlation Co-efficient | P Value |
| Total IC | NA | | 0.9 | <.0001 |
| Overall PD-1 IC | NA | | 0.8 | <.0001 |

**Mixed PD-L1 expression: Constitutive +Adaptive**

Diffuse (N=4)

| | Tumor PD-L1 expression | | Immune cell PD-L1 expression | |
|---|---|---|---|---|
| Immune cells | Correlation Co-efficient | P Value | Correlation Co-efficient | P Value |
| Total IC | -0.3 | 0.73 | -0.3 | 0.74 |
| Overall PD-1 IC | -0.6 | 0.37 | 0.8 | 0.2 |

4 SCC’s lacked PD-L1 expression in tumor cells and immune cells

### Table S3: Comparison of TPS, CPS with clinical characteristics of SCC participants (N=52)

| Patient Characteristics | | TPS+ | TPS- | CPS+ | CPS- | TPS | CPS |
|---|---|---|---|---|---|---|---|
| | | N=27 | N=25 | N=48 | N=4 | Fisher's exact P-Value | Fisher's exact P-Value |
| Sex | Female | 12 (44.4%) | 7 (28%) | 18 (37.5%) | 1 (25%) | 0.26 | 1 |
| | Male | 15 (55.6%) | 18 (72%) | 30 (62.5%) | 3 (75%) | | |
| Age | 25-49 | 4 (14.8%) | 5 (20%) | 9(18.8%) | 0 | 0.91 | 0.72 |
| | 50-69 | 20 (74.1%) | 18 (72%) | 34(70.8%) | 4 (100 %) | | |
| | 70+ | 3 (11.1%) | 2 (8%) | 5 (10.4%) | 0 | | |
| Race | Black | 3 (11.1%) | 3 (12%) | 5 (10.4%) | 1 (25%) | 0.69 | 0.73 |
| | Hispanic | 1 (3.7%) | 3 (12%) | 4 (8.3%) | 0 | | |
| | Mixed | 0 | 1 (4%) | 1 (2.1%) | 0 | | |
| | Unknown | 2 (7.4%) | 1 (4%) | 3 (6.3%) | 0 | | |
| | White | 21 (51.9%) | 17 (68%) | 35 (72.9%) | 3 (75%) | | |
| Grade | Moderately | 10 (37%) | 12 (48%) | 20 (41.7%) | 2 (50%) | 0.19 | 0.19 |
| | Poor | 4 (14.8%) | 7 (28%) | 9 (18.8%) | 2 (50%) | | |
| | Well | 13 (48 % | 6 (24%) | 19 (39.6%) | 0 | | |
| Stage | I | 8 (29.6%) | 9 (36%) | 17 (35.4%) | 0 | 0.70 | 0.27 |
| | II | 14 (51.9%) | 8 (32%) | 20 (41.7%) | 2 (50%) | | |
| | III-IV | 3 (11.1%) | 7 (28%) | 8 (16.7%) | 2 (50%) | | |
| Immune Status | HIV-, non-immunosuppressed | 14 (60.9%) | 9 (39.1%) | 22 (95.7%) | 1 (4.3%) | 0.2 | 0.67 |
| | HIV -, immunosuppressed | 0 | 2 (100%) | 2 (100%) | 0 | | |
| | PLWH | 12 (46.2%) | 14 (53.8%) | 23 (88.5%) | 3 (11.5%) | | |

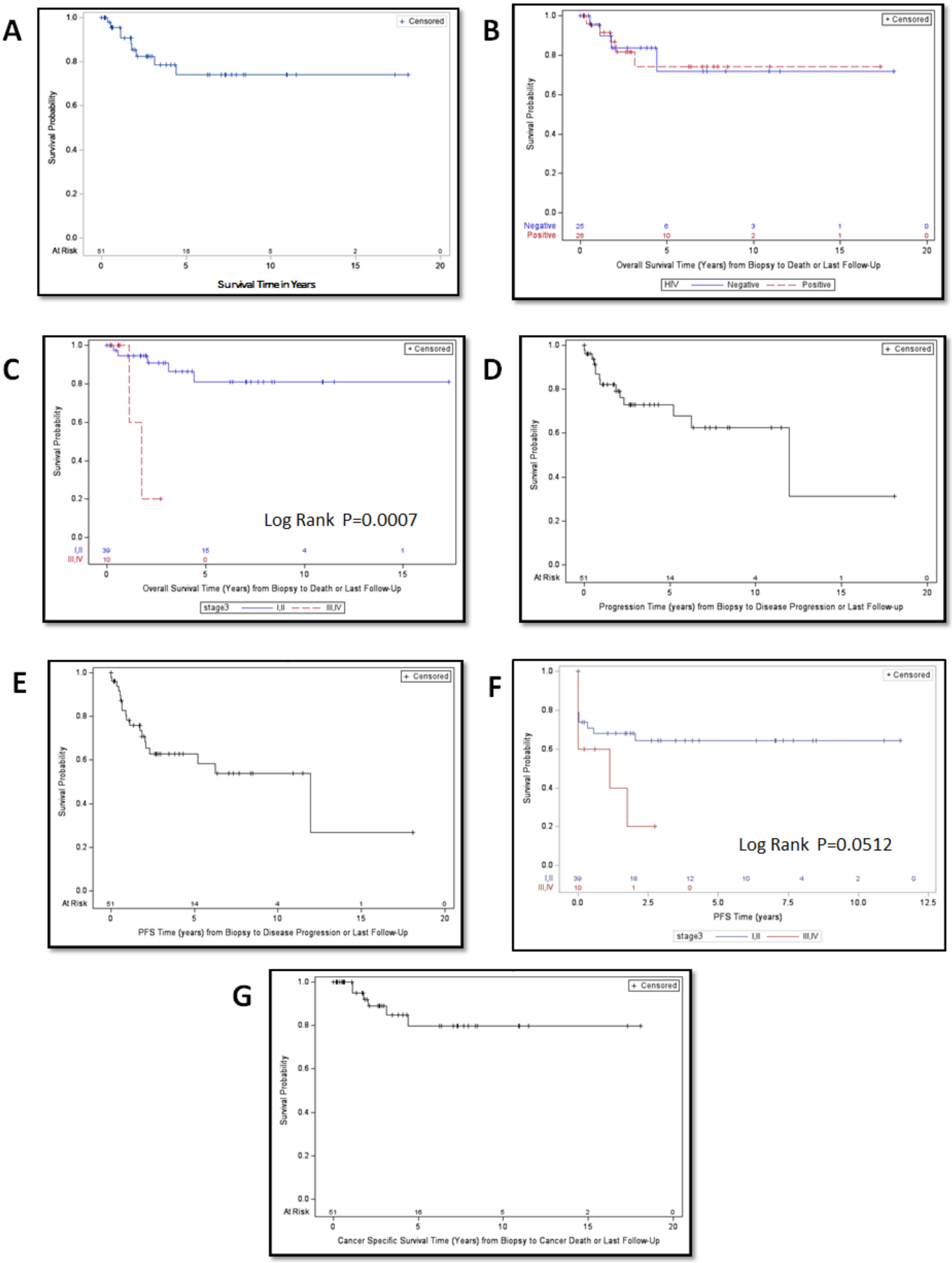
A
+ Censored
Survival Probability
At Risk
51 16 5 2 0
Survival Time in Years
B
+ Censored
Survival Probability
Negative 25 6 3 1 0
Positive 26 10 2 1 0
Overall Survival Time (Years) from Biopsy to Death or Last Follow-Up
HIV Negative Positive
C
+ Censored
Survival Probability
Log Rank P=0.0007
I,II 39 15 4 1
III,IV 10 0
Overall Survival Time (Years) from Biopsy to Death or Last Follow-Up
stage3 I,II III,IV
D
+ Censored
Survival Probability
At Risk 51 14 4 1 0
Progression Time (years) from Biopsy to Disease Progression or Last Follow-up
E
+ Censored
Survival Probability
At Risk 51 14 4 1 0
PFS Time (years) from Biopsy to Disease Progression or Last Follow-Up
F
+ Censored
Survival Probability
Log Rank P=0.0512
I,II 39 18 12 10 4 2 0
III,IV 10 1 0
PFS Time (years)
stage3 I,II III,IV
G
+ Censored
Survival Probability
At Risk 51 16 5 2 0
Cancer Specific Survival Time (Years) from Biopsy to Cancer Death or Last Follow-Up

**Figure S1:** Kaplan-Meier analyses of anal SCC participants. (a) overall survival (OS)(b) overall survival stratified by HIV status (log-rank p=0.97) (c) overall survival stratified by tumor stage (log-rank p=0.0007). (d) disease progression (e) progression-free survival (PFS) (f) Progression-free survival stratified by tumor stage (p=0.051) (g) Cancer-Specific Survival (CSS)

### Table S4(a): Cox proportional hazards model of Progression-free survival

| Model | Variable | Categories | Events (%) | Hazard Ratio | Lower 95% CI | Upper 95% CI | P-value |
|---|---|---|---|---|---|---|---|
| 2 | CPS | 0.0-4.5 | 6/13 (46.2%) | 5.8 | 0.70 | 48 | 0.10 |
| | | 4.5-8.1 | 5/13 (38.5%) | 5.9 | 0.69 | 51 | 0.10 |
| | | 8.1-20.0 | 7/13 (53.8%) | 8.0 | 0.99 | 65 | 0.05 |
| | | > 20 | 1/12 (8.3%) | 1.00 | | | |

### Table S4(b): Cox proportional hazards model of Progression-free survival

| Model | CPS | Events (%) | Hazard Ratio | Lower 95% CI | Upper 95% CI | P-Value |
|---|---|---|---|---|---|---|
| 4 | Negative | 3/4 (75.0%) | 1.96 | 0.57 | 6.8 | 0.28 |
| | Positive | 16/47 (34.0%) | 1.00 | | | |

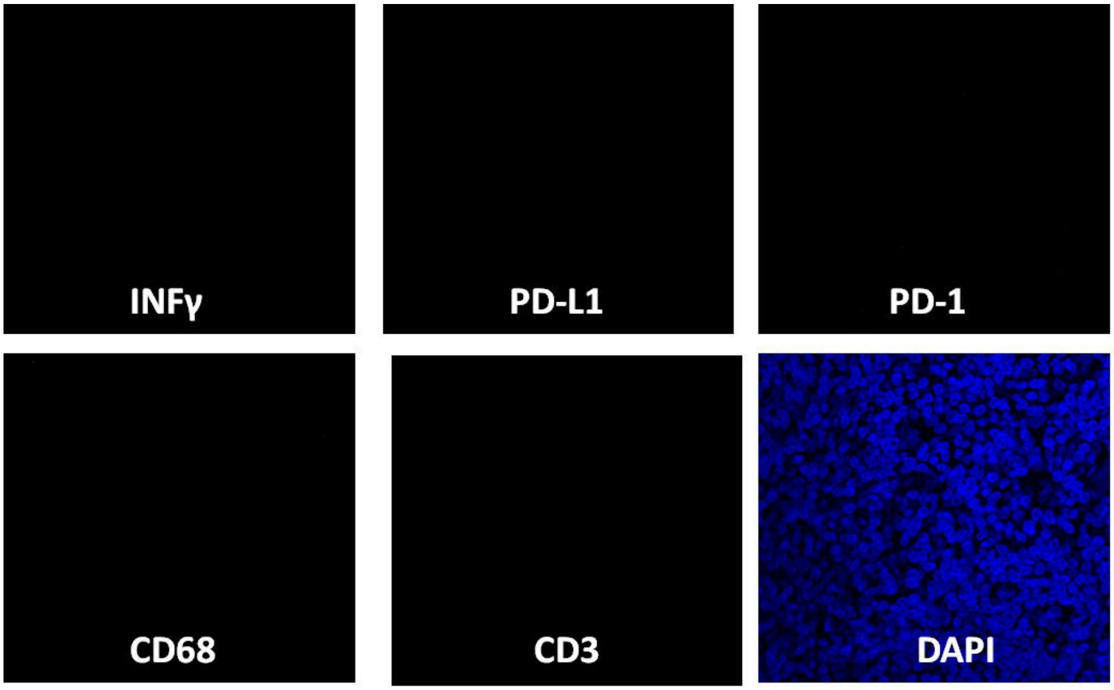


**Figure S2:** In situ hybridization and multiplex immunofluorescence staining of tonsil tissue. Control probes were used for IFN-γ (RNAscope probe, pink), PD-L1 (RNAscope probe, yellow), PD-1 (RNAscope probe, blue), and CD68 (RNAscope probe, green); CD3 (immunohistochemistry, no primary antibody). Counterstaining with DAPI [63X, N=1].
